# Regional Differences in the US in Outcomes with Preeclampsia

**DOI:** 10.1101/2024.05.20.24307230

**Authors:** Aardra Rajendran, Yisi Liu, Theresa M. Boyer, Arthur J. Vaught, Allison G. Hays, Josef Coresh, Dhananjay Vaidya, Erin D. Michos, Anum S. Minhas

**Affiliations:** Department of Internal Medicine, Johns Hopkins University School of Medicine, Baltimore, MD, USA; Department of Pediatrics, Johns Hopkins University School of Medicine, Baltimore, MD, USA; Department of Epidemiology, Johns Hopkins Bloomberg School of Public Health, Baltimore, MD, USA; Division of Maternal Fetal Medicine, Department of Gynecology and Obstetrics, Johns Hopkins University, Baltimore, Maryland; Division of Acute Care and Adult Trauma Surgery, Department of Surgery, Johns Hopkins University, Baltimore, Maryland; Division of Cardiology, Department of Medicine, Johns Hopkins University School of Medicine, Baltimore, MD, USA; Ciccarone Center for the Prevention of Cardiovascular Disease, Division of Cardiology, Johns Hopkins University School of Medicine, Baltimore, MD, USA; Division of General Internal Medicine, Department of Medicine, Johns Hopkins University School of Medicine, Baltimore, MD, USA

**Author notes:** **Address for Correspondence:** Aardra Rajendran, MD, 1800 Orleans Street, Carnegie 591A, Baltimore, MD 21287, Twitter: @AardraRajendran. **Disclosures:**.

## Abstract

This study explores preeclampsia outcomes across US regions and examines regional differences in specific preeclampsia-associated pregnancy complications and disease management. Patient-reported measures were obtained from The Preeclampsia Registry, an open-access database composed of women with at least one pregnancy diagnosed with a hypertensive disorder of pregnancy. Pregnancies and associated outcomes were stratified by US region (Northeast, Midwest, South and West). Among 2,667 pregnancies of which 92% were in White women, maximum systolic blood pressure at any time in pregnancy was highest among women in the South and Midwest (p=0.039). Furthermore, more women in the South received pre-pregnancy anti- hypertensives (p=0.026) and antenatal steroids (p=0.025) and delivered at an earlier gestational age (p=0.014) compared to women in other regions. Pregnancy complications such as elevated liver enzymes were higher in women in the South (p=0.019), and women in the South and West had additional end-organ damage such as renal complications (p<0.001) and hemolysis (p=0.008) as compared to women in other regions. Further investigation is needed to assess whether healthcare access or policy could be contributing to these regional discrepancies.

## Introduction

Preeclampsia affects 2-8% of all pregnancies and is associated with adverse maternal-fetal outcomes and increased cardiovascular disease risk. Studies have demonstrated regional geographic differences in the prevalence of preeclampsia in the United States (US). Women in the South are at higher risk for developing preeclampsia and have an increased risk of maternal mortality compared to women in the Northeast (1,2). In this study, we aim to gain a deeper understanding of preeclampsia outcomes across US regions and examine regional differences in specific preeclampsia-associated pregnancy complications and disease management.

## Methods

Retrospective deidentified patient-reported measures were obtained from The Preeclampsia Registry (TPR), an open-access database composed of women with at least one pregnancy diagnosed with a hypertensive disorder of pregnancy (HDP) (chronic hypertension, gestational hypertension, preeclampsia, eclampsia, or chronic hypertension with superimposed preeclampsia or eclampsia). Participants self-reported their diagnosis of HDP and personal, family, medical, and pregnancy history, including clinical measures and laboratory data through an online questionnaire. Elevated liver enzymes, renal complications, hemolysis, and thrombocytopenia were defined by standard of care laboratory measurements. Self-reported outcomes in TPR were obtained using well-validated instruments (data harmonization criteria established by Co-Lab, obstetric data definitions validated by ACOG, and common data points suggested by the Women’s Health Registries Alliance)(3). Approximately 40% of patients self-enrolled after finding TPR through an internet search and an additional 25% were recruited through foundation events. Greater than 90% of pregnancies occurred after 2003 and <1% occurred before 1984, thereby ensuring that the majority of pregnancies and their management reflect current health practices. Patients were asked to provide annual updates following registry inclusion; however, follow-up data was not analyzed given limited availability. In this study, only pregnancies with preeclampsia were included in analysis and data were grouped by US region as defined by the US Census: Northeast, Midwest, South and West. Clinical outcomes were compared across regions using Chi-square test.

## Results

Among 2,667 pregnancies, mean maternal age at delivery was 29.6+5.2 years and 92% of women were White (Table 1). Maximum systolic blood pressure at any time during pregnancy was higher in women in the South (185[170-201] mmHg) and Midwest (185[168-202] mmHg) compared to women in the Northeast (182.5[160-200] mmHg) and the West (180[165-200] mmHg, p=0.039). Prior to pregnancy, more women in the South were taking anti-hypertensive medications (16.8%) compared to women in the Northeast (7.5%, p=0.026). During pregnancy, prevalence of anti-hypertensive use was higher in the South (31.3%) compared to the Northeast (24.6%), although the difference did not reach statistical significance. Women in the South had the earliest gestational age at delivery (33.8+4.7 weeks) while women in the Northeast had the latest gestational age (34.5+4.3 weeks, p=0.014), and furthermore, more women in the South required antenatal steroids for preterm delivery (12.3%) compared to women in the Northeast (7.9%, p=0.025). Preeclampsia-associated pregnancy complications, including elevated liver enzymes, renal complications, and hemolysis, were generally higher among women in the West and South as compared to women in the Midwest and Northeast. There were no significant differences in the rates of Caesarean-section across regions (68.8% Northeast; 67.4% South).

**Table 1.**
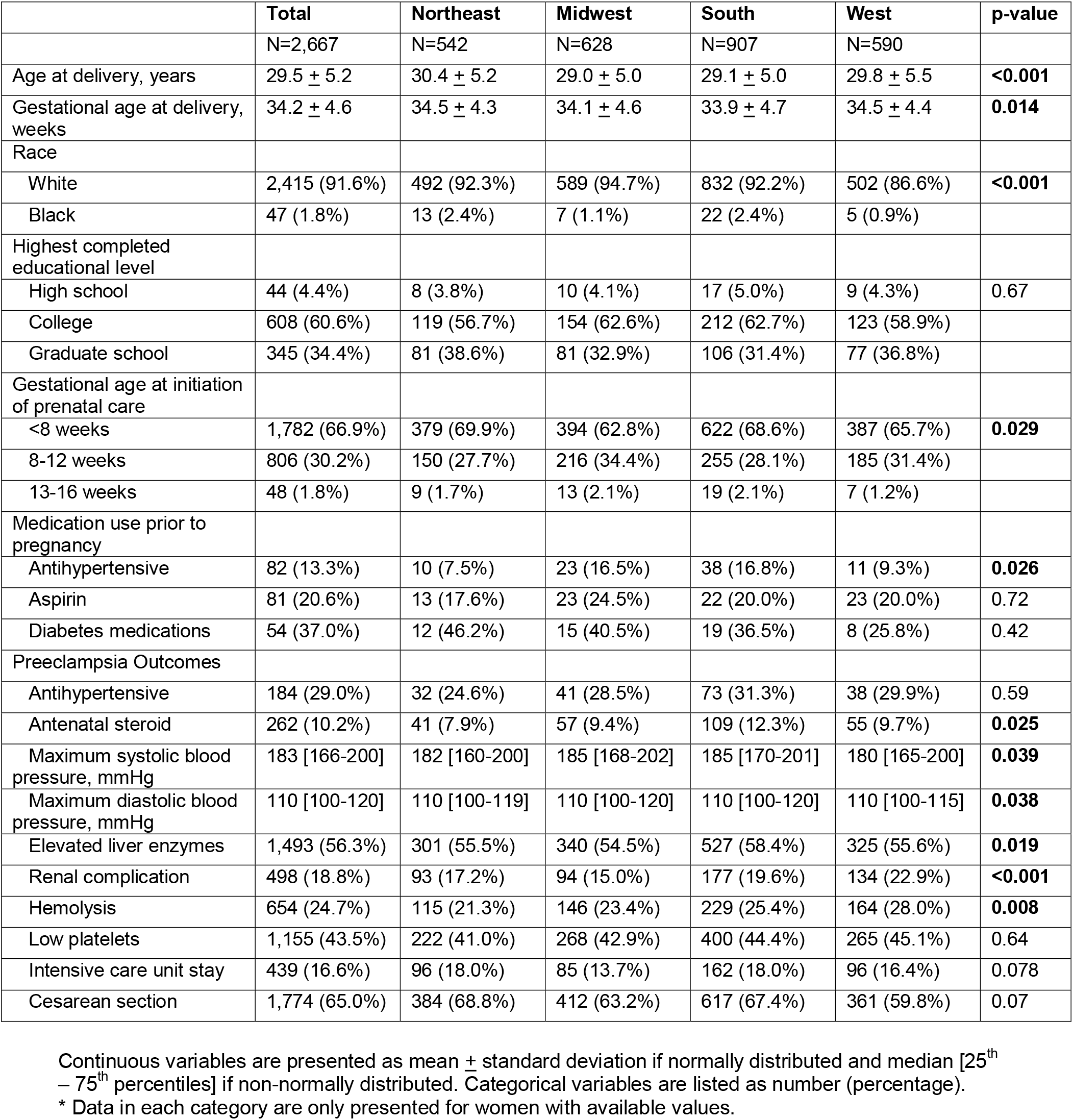
Preeclampsia Participant Demographics and Clinical Outcomes, Stratified by US Census Region.

## Discussion

This study suggests that preeclampsia-associated pregnancy complications may be greater among women in the South, thus building on prior findings that women in the South are at higher risk of developing preeclampsia compared to women in the Northeast (1). Women were more likely to require anti-hypertensive medications prior to pregnancy and had higher maximum blood pressure during pregnancy compared to women in the Northeast. They also delivered at an earlier gestational age and had increased requirements for antenatal steroid use. Lastly, women in the South and West had more preeclampsia-associated end-organ damage as compared to women in the Northeast and Midwest.

These regional differences could potentially be attributable to differences in preeclampsia risk factors, demographic factors, and/or socioeconomic disparities related to healthcare access. BMI data was unavailable in this dataset; however, obesity is an important risk factor for the development of preeclampsia and its prevalence has been increasing over the years, particularly in the South (4). Furthermore, obesity rates are highest in non-Hispanic Black women, and Black women independently have higher incidences of preeclampsia and preeclampsia-associated complications (5). Increased adverse pregnancy outcomes in the South may also be due to decreased prenatal care and birthing facilities (2).

This study represents data from across the US and is not limited by institutional or targeted recruitment. Importantly, it includes patient-reported outcomes; however, as participation is voluntary, selection bias is possible. Additionally, the majority of women enrolled are White, and analyses between regional differences and race/ethnicity was not performed due to sample size limitations. Nonetheless, we find differences in key clinical outcomes and variables by geographic region in the US. Future studies should be performed to investigate whether variations in healthcare access or policy could explain regional disparities.

## Data Availability

All data was obtained from The Preeclampsia Registry through Preeclampsia Foundation.

## Acknowledgements

The authors are grateful to the women who chose to participate in the Preeclampsia Registry and to the Preeclampsia Foundation for providing the data.

